# Scoping Review of Existing Stroke Guidelines; Argument for a Value-Added Change

**DOI:** 10.1101/2021.02.12.21251660

**Authors:** Tissa Wijeratne, Carmela Sales, Mihajlo (Michael) Jakovljevic, Leila Karimi

## Abstract

**Background and Purpose:** Stroke represents one of the most important causes of morbidity ( eighty million patients with disabling of ongoing effects of stroke at a given time, globally) and mortality (the second leading cause of death) worldwide. Innovative systems biology-based approach is likely to increase the understanding of the underpinning of acute stroke promise to enhance stroke prevention, acute treatment, and neurorehabilitation. Recent growing body of evidence with shared pathobiology with COVID-19 and the critically important role of inflammation in the context of stroke points to far-reaching consequences of acute stroke, just as in the case of COVID-19 ( post-acute event issues as well as long term issues).

So far, stroke typically defined by late-appearing disease manifestation by the range of stroke subtypes as defined by the WHO or American Stroke Association. This definition neglects the underlying pathobiological mechanisms such as low-grade chronic inflammation and already compromised vascular system. Diseases such as stroke is hardly a simple result of a single problem, but rather a complex cascade of pathobiological processes and interactions in a complex biochemical environment. The evidence of changes in innate immunity and adaptive immunity during the index event of acute stroke and recovery over next 3-12 months can be easily elicited with simple bedside blood tests such as neutrophil-lymphocyte ratio (NLR) with well over 300 published papers including several systematic reviews and meta-analyses confirming this. Global standard operating procedures (SOP) of stroke care dictated by the national and international stroke guidelines at present. It is imperative to explore the evidence of systems biology approach in current stroke guidelines. This is likely to be a key turning point in managing stroke across the continuum (prevention, management of acute event and rehabilitation).

**Methods:** We systematically searched for guideline recommendation on the day-to-day use of peripheral inflammatory markers such as NLR published in the English language between January 1, 2005, and December 2020. Any other evidence of systems biology-based approach or recommendation was explored within the selected guidelines for this scoping review. Only the latest guideline per writing group was selected. Each guideline was analyzed independently by 2 to 4 authors to determine clinical scenarios explained/given, scientific evidence used, and recommendations presented in the context of systems biology.

**Results:** The scoping review found 3,830 (3830) titles with 119 guidelines from 46 countries included for this review ( Figure 1; PRISMA diagram). Stroke-related organizations wrote Sixty-five per cent of the guidelines while national ministries wrote a fewer number of guidelines. We were primarily interested in recommendations for acute management in AIS published in the English language. Fifteen eligible guidelines were identified from 15 different countries/regions. None of the guidelines recommended the routine use of peripheral markers of inflammation, such as NLR, among their acute assessment and management recommendations. None of the existing guidelines explored the systems biology approach to one of the most complex diseases affecting the human brain, stroke.

Figure 1
Acute Ischemic Stroke Guidelines Worldwide

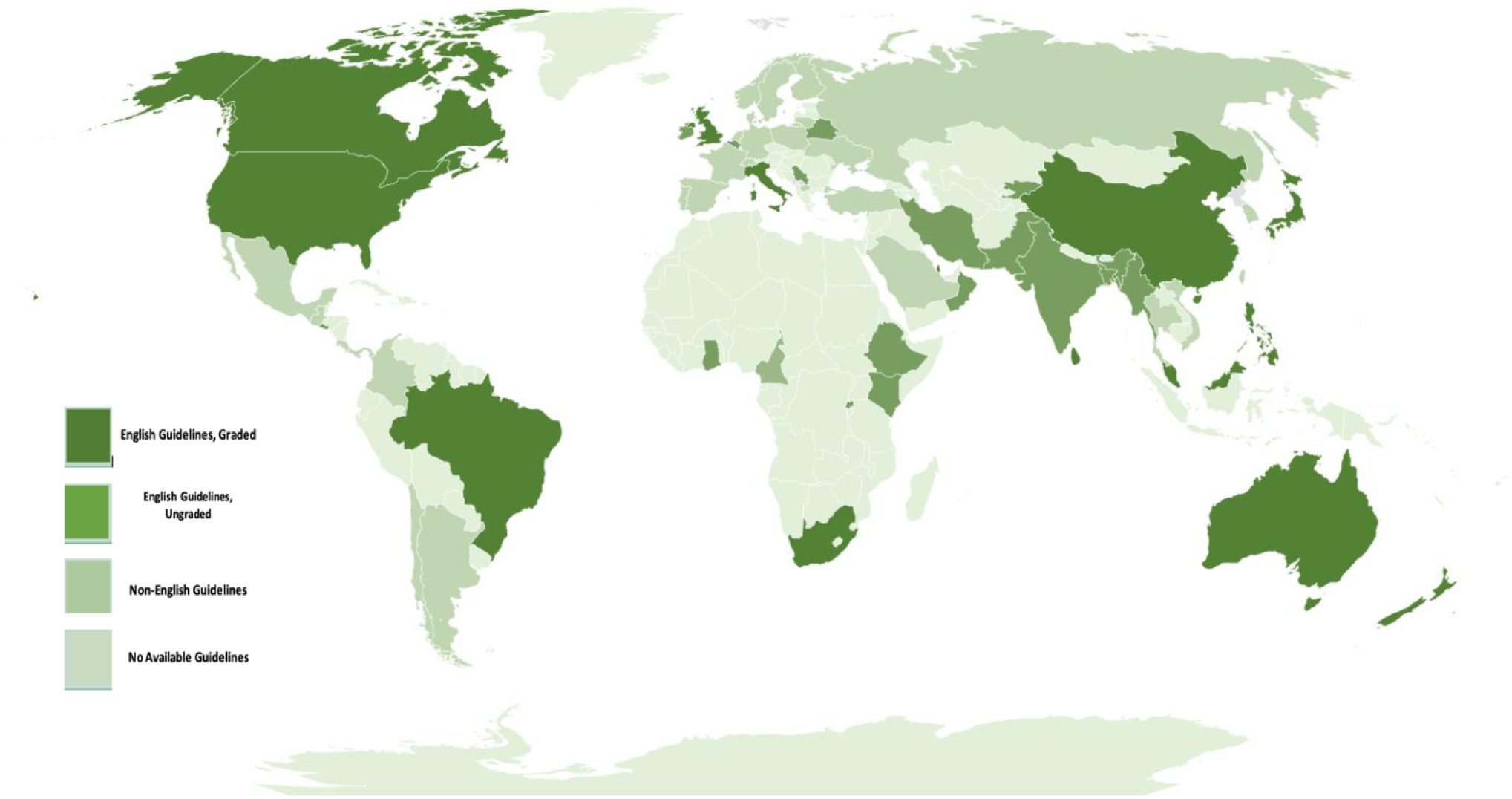

Figure 2:
PRISMA Diagram

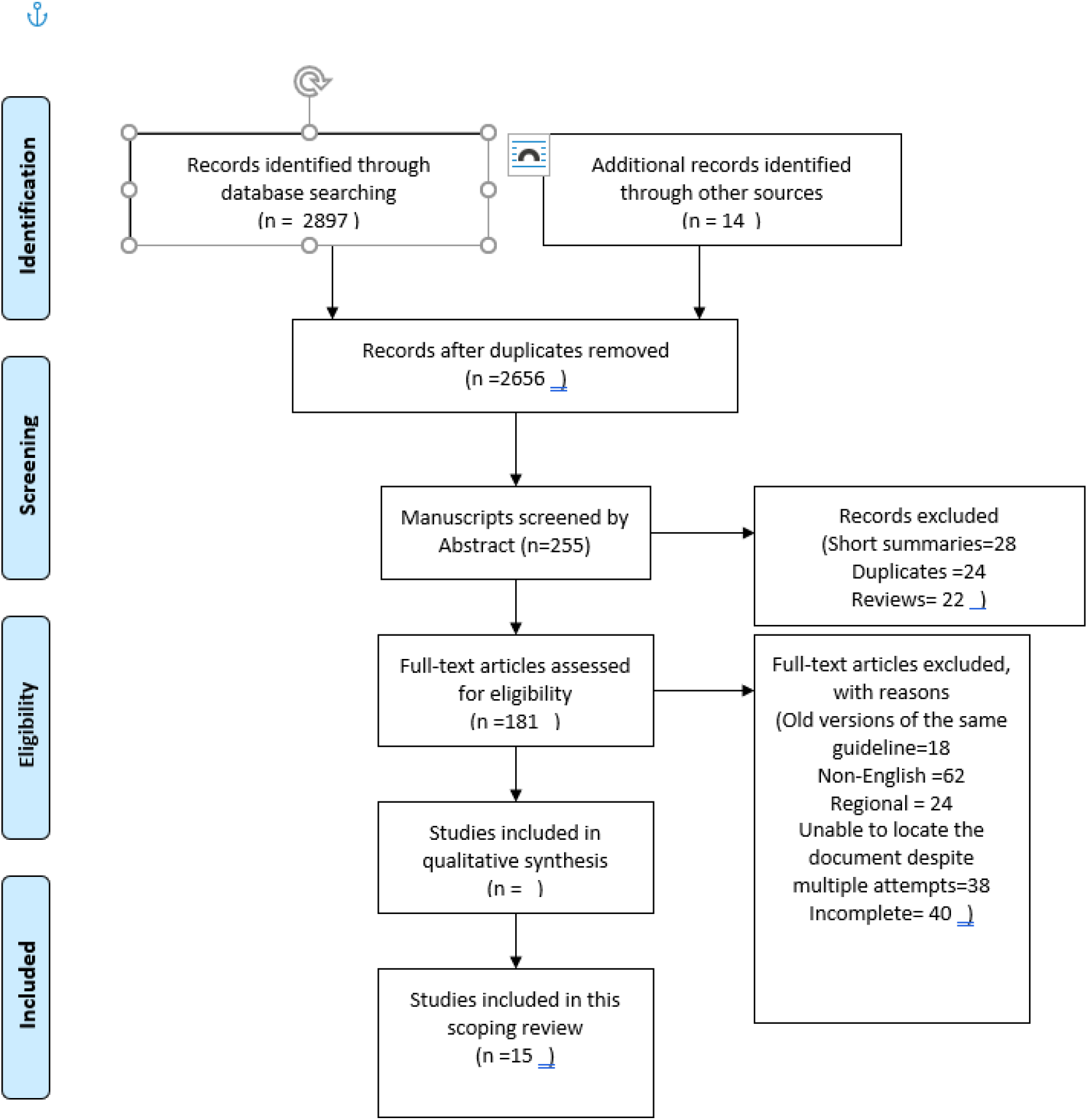

**Conclusions:** This systematic review has identified a significant evidence-practice gap in all existing national stroke guidelines published in English medium as of October 2020. These guidelines included the only current “living stroke guidelines, Stroke Guidelines from Australia with a real opportunity to modernize the living stroke guidelines with systems biology approach and provide 2020 vision towards better stroke care globally.

Investigation of complex disease such as stroke is best served through a systems biology approach. One of the easiest places to start is simple blood tests such as total white cell count and NLR. Systems biology approach point us towards simple tools such immune-inflammatory index (SII), Sunshine Prognostic Score (SPS) which should pave the way for the stroke physician community address the challenges in systems biology approach in stroke care. These challenges include translating bench research to the bedside, managing big data ( continuous pulse, blood pressure, sleep, Oxygen saturation, progressive changes in NLR, SII, SPS, etc.). Working with an interdisciplinary team is also provide a distinct advantage.

## Introduction

Evidence-based medicine calls for the utilization of widely available clinical guidelines especially for the management of common conditions which have an impact on mortality and morbidity such as acute ischemic stroke (AIS). The first of this kind was published in 1974 which was entitled ‘Prologue to Guidelines for Stroke Care’, a compendium of articles compiled by Neurologists on the management of cerebrovascular disease (1) It was not until more than 20 years later that the Cochrane Collaboration Stroke Review Group convened and initiated the task of constructing a systematic guideline for the management of acute stroke (2).

Clinical guidelines are essential tools to improve the quality of healthcare systems. Noncommunicable diseases (NCDs) and vascular ones in particular are lifetime complex chronic conditions, which are expensive to treat(3) and expose tremendous budget impact affecting entire financial sustainability of contemporary health systems (4). Factors which are crucial for a clinical guideline to be successfully crafted is team collaboration and multidisciplinary engagement (5, 6). Furthermore, these should be tailor-fitted to individual country needs, hence, the non-existence of a universally implemented guideline (6). The use of tools to assess the quality of evidence also aids clinicians to interpret the recommendations according to the weight of evidence (7). Potential barriers to non-adherence include unfamiliarity, lack of agreement and outcome expectancy, as well as the significant impact of the precedent guideline (8).

Perhaps one of the game-changers in the history of medicine is the development of clinical guidelines for the management of AIS. The wealth of data from clinical trials on reperfusion therapies paved the way for the American Heart Association (AHA) and the Canadian Stroke Consortium to publish their respective recommendations on the acute intervention of cerebrovascular ischemia(9, 10). Through time, various versions of clinical guidelines have also been published in different languages with the primary objective of implement ability according to the resources available in each country. While constructs behind these standard procedures are anchored on the same theory, some degree of variability still occurs(11). To date, there are no studies which specifically look at the differences in the clinical guidelines on acute ischemic stroke in a worldwide. It is in this light that this study was conceived by the first author (TW).

## Methodology

The authors of this review used the Arksey and O’Malley methodology to identify and extract useful literature. The steps undertaken include (1) research question identification; (2) relevant literature identification; (3) screening and selection of relevant literature;(4) data charting; and (5) analyzing, summarizing, and reporting results.

MEDLINE, Cochrane and CINHAL databases were searched to identify useful keywords. Subsequently, the identified keywords were used to search the same databases for relevant studies. Literature were first screened at the title, and the abstract level then the full text articles.

Following search term were employed based on the PICO strategy. Topic= “ country name” AND TOPIC=“guideline” OR “ clinical protocols” OR “recommendations” OR “standards” AND TOPIC = stroke OR cerebrovascular disorder OR cerebrovascular accident.

Guideline repositories such as the National Guideline Clearinghouse, the Scottish Intercollegiate Guidelines Network (SIGN) and Professional stroke societies were also searched. Individual bibliographies were also manually searched. Studies were included if they met the following criteria: a) published after year 2000 b) guidelines on stroke and/or post-stroke rehabilitation c) graded recommendations d) written in English. Titles and abstracts were initially screened (TW) and any full-text articles were further appraised (TW, CS). Any disagreement was adjudicated by an independent reviewer (LK). Guidelines which were updated in a modular format and published over separate papers were treated as one guideline.

## Results

### Guideline characteristics

Figure 1 shows the diagram on available stroke guidelines worldwide. Majority of the countries have no available published national guidelines while a number have guidelines but no graded recommendations. A significant majority also have guidelines published in their own language while 14 countries have their own published, graded, English clinical guidelines, with the one from the European Stroke Organization as a separate entity.

A total of 2897 titles were identified in the electronic search. Fourteen additional records were identified through other sources. After removal of duplicates and screening at the title level, 255 articles were further reviewed at the abstract level. Hundred and eighty-one papers were thoroughly assessed by the two authors (TW and CS) for eligibility. A total of 15 guidelines were included in this scoping review.

Table 1 outlines the characteristics of the 15 clinical guidelines included in this study while Table 2 describes the recommendations of individual guidelines and the specific grades tagged on each guideline parameter.

**Table 1.**
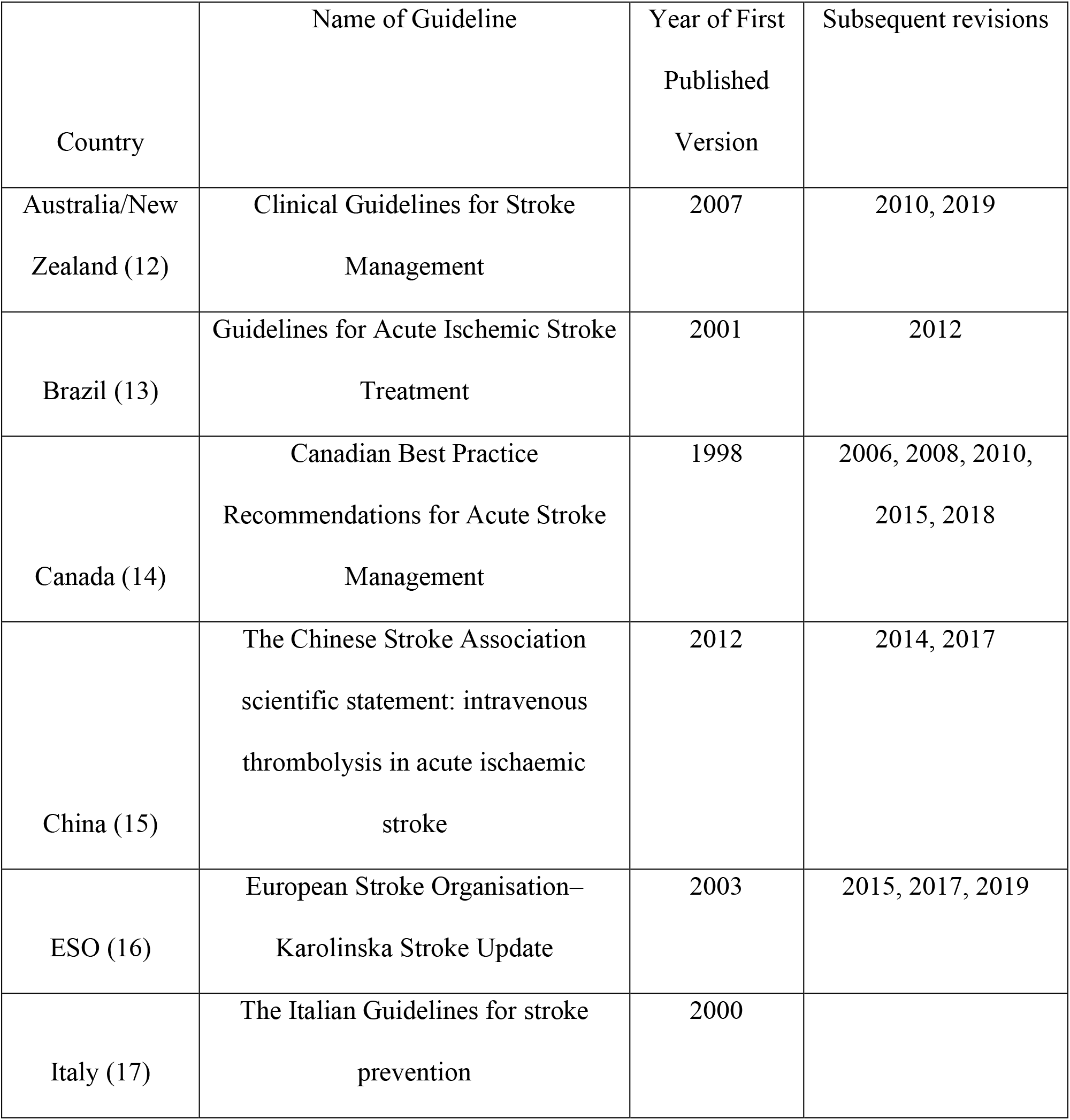
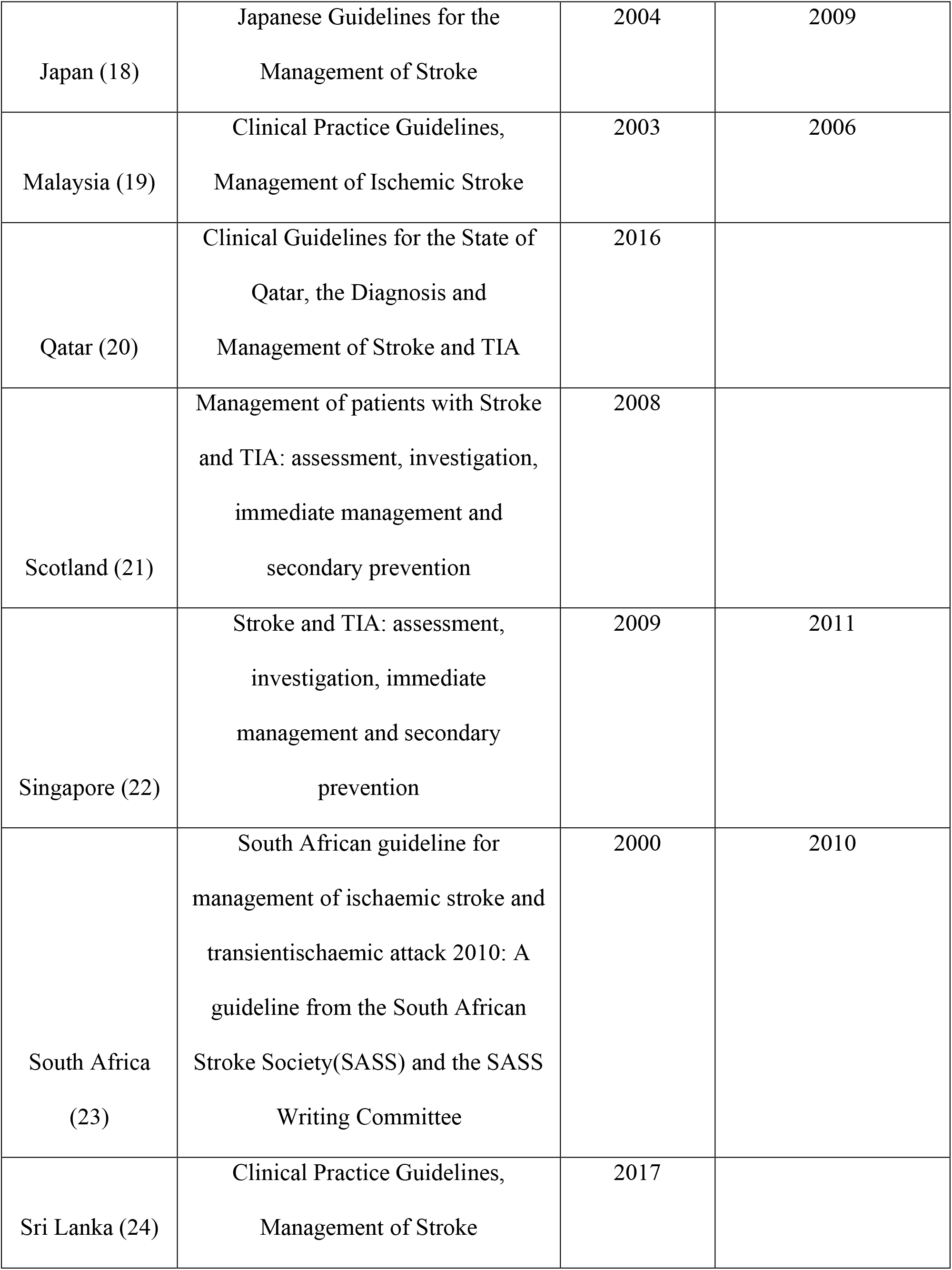
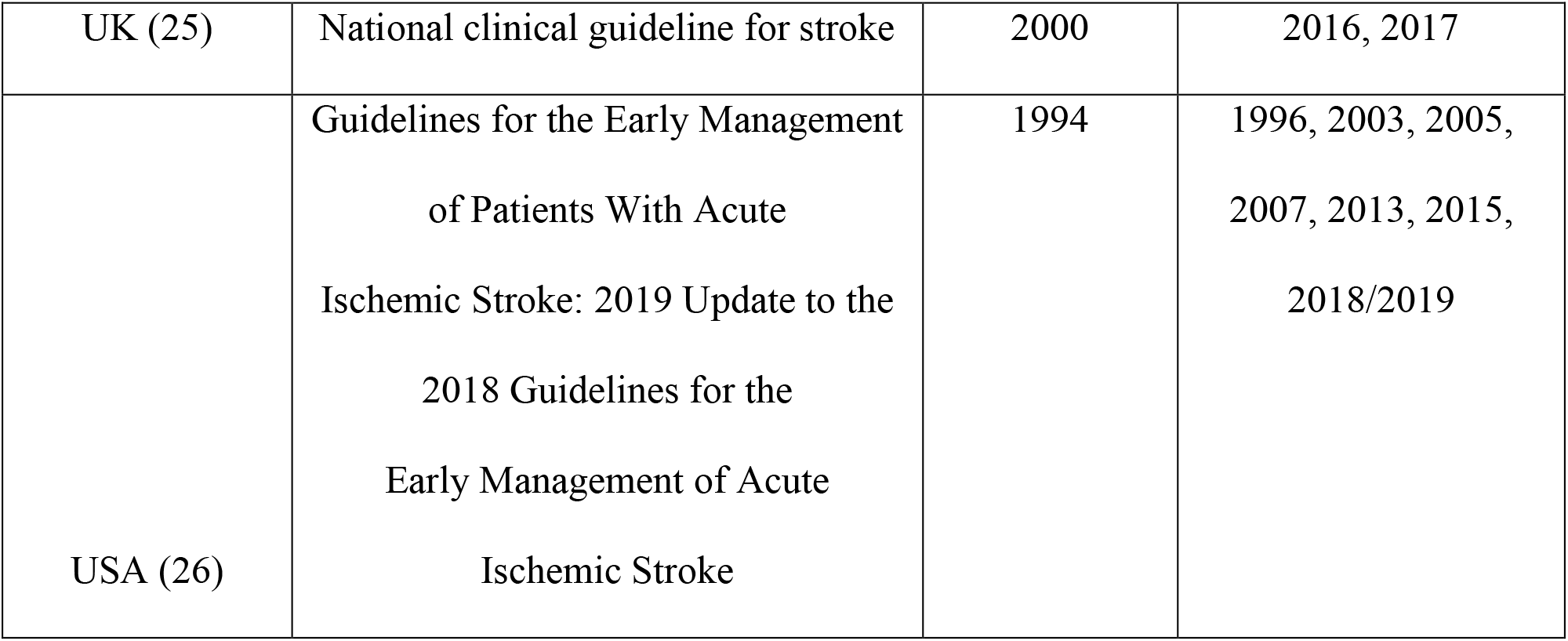
Summary of Acute Ischemic Stroke Clinical Guidelines

| Country | Name of Guideline | Year of First Published Version | Subsequent revisions |
| --- | --- | --- | --- |
| Australia/New Zealand (12) | Clinical Guidelines for Stroke Management | 2007 | 2010, 2019 |
| Brazil (13) | Guidelines for Acute Ischemic Stroke Treatment | 2001 | 2012 |
| Canada (14) | Canadian Best Practice Recommendations for Acute Stroke Management | 1998 | 2006, 2008, 2010, 2015, 2018 |
| China (15) | The Chinese Stroke Association scientific statement: intravenous thrombolysis in acute ischaemic stroke | 2012 | 2014, 2017 |
| ESO (16) | European Stroke Organisation–Karolinska Stroke Update | 2003 | 2015, 2017, 2019 |
| Italy (17) | The Italian Guidelines for stroke prevention | 2000 |  |
| Japan (18) | Japanese Guidelines for the Management of Stroke | 2004 | 2009 |
| Malaysia (19) | Clinical Practice Guidelines, Management of Ischemic Stroke | 2003 | 2006 |
| Qatar (20) | Clinical Guidelines for the State of Qatar, the Diagnosis and Management of Stroke and TIA | 2016 |  |
| Scotland (21) | Management of patients with Stroke and TIA: assessment, investigation, immediate management and secondary prevention | 2008 |  |
| Singapore (22) | Stroke and TIA: assessment, investigation, immediate management and secondary prevention | 2009 | 2011 |
| South Africa (23) | South African guideline for management of ischaemic stroke and transient ischaemic attack 2010: A guideline from the South African Stroke Society(SASS) and the SASS Writing Committee | 2000 | 2010 |
| Sri Lanka (24) | Clinical Practice Guidelines, Management of Stroke | 2017 |  |
| UK (25) | National clinical guideline for stroke | 2000 | 2016, 2017 |
| USA (26) | Guidelines for the Early Management of Patients With Acute Ischemic Stroke: 2019 Update to the 2018 Guidelines for the Early Management of Acute Ischemic Stroke | 1994 | 1996, 2003, 2005, 2007, 2013, 2015, 2018/2019 |

**Table 2.**
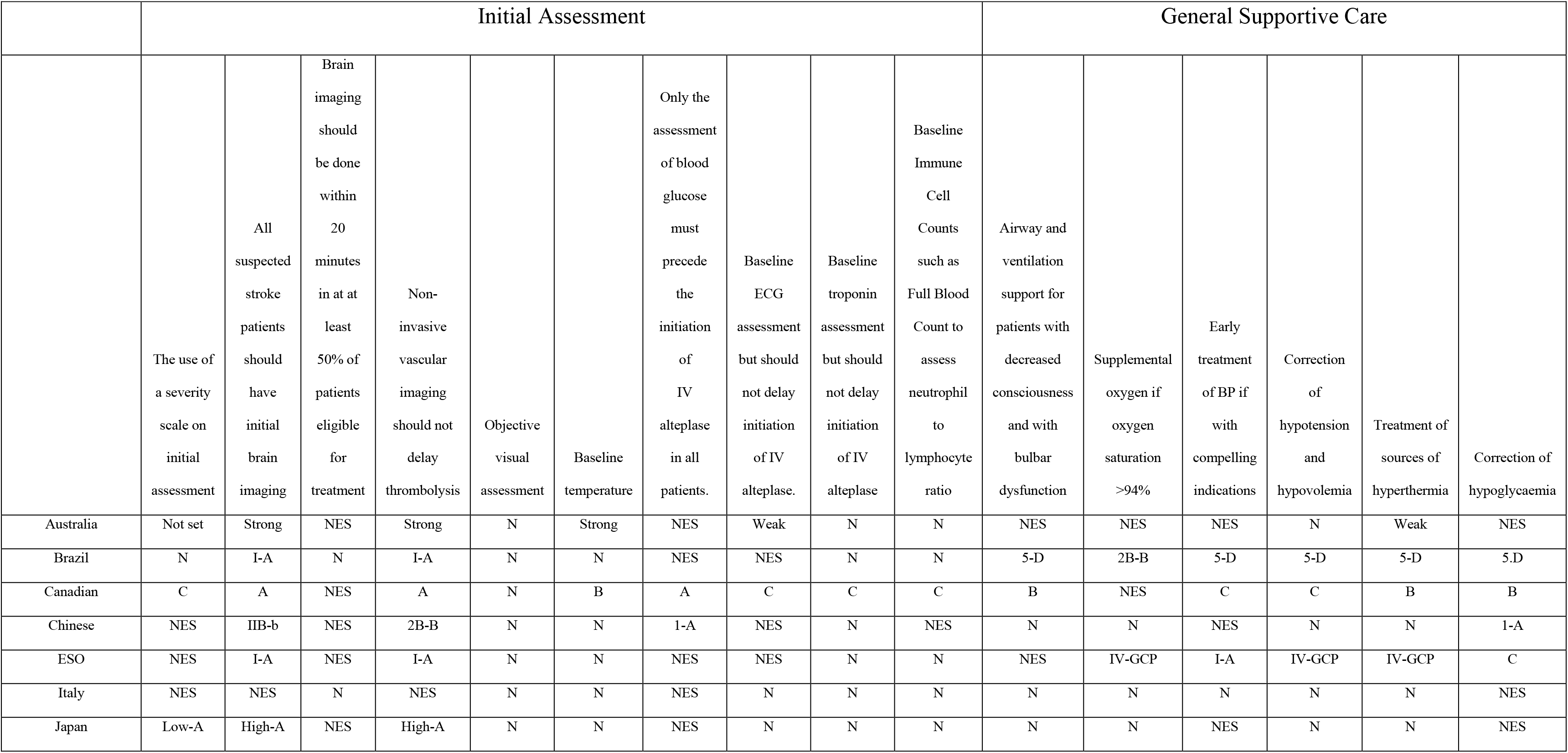
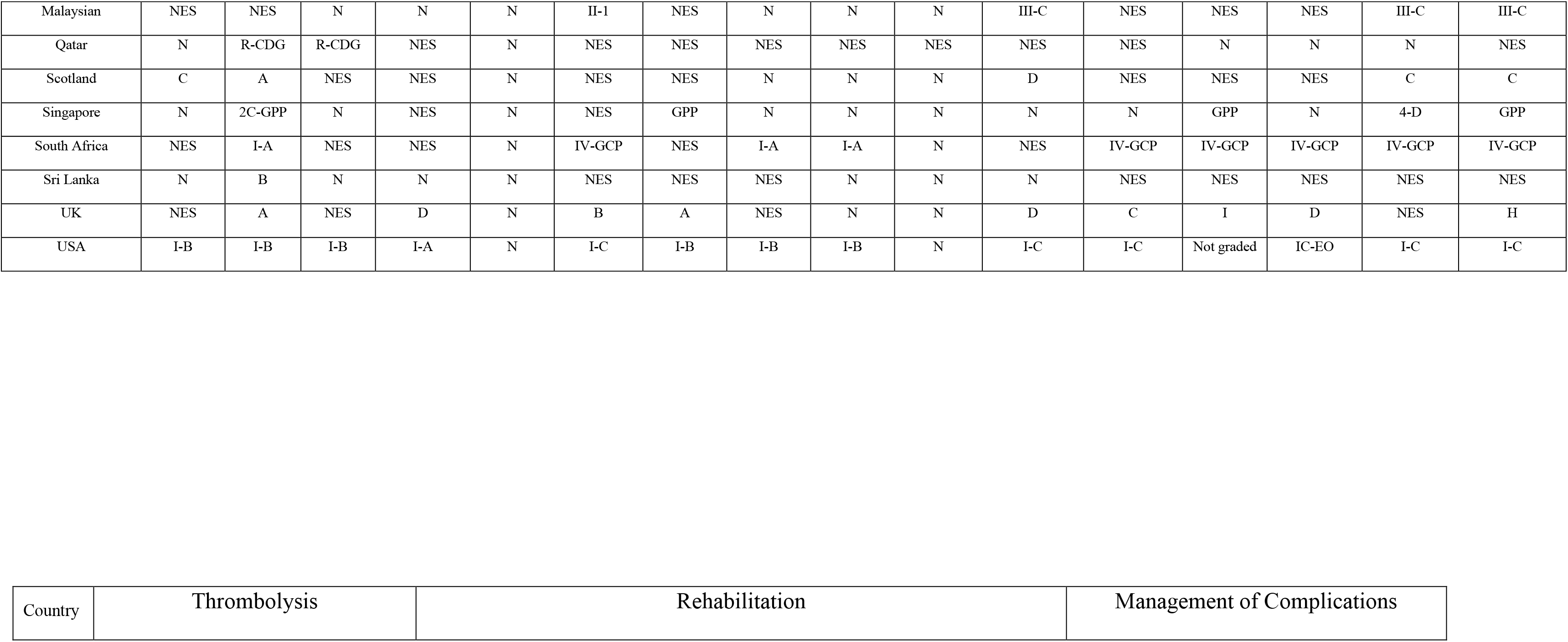
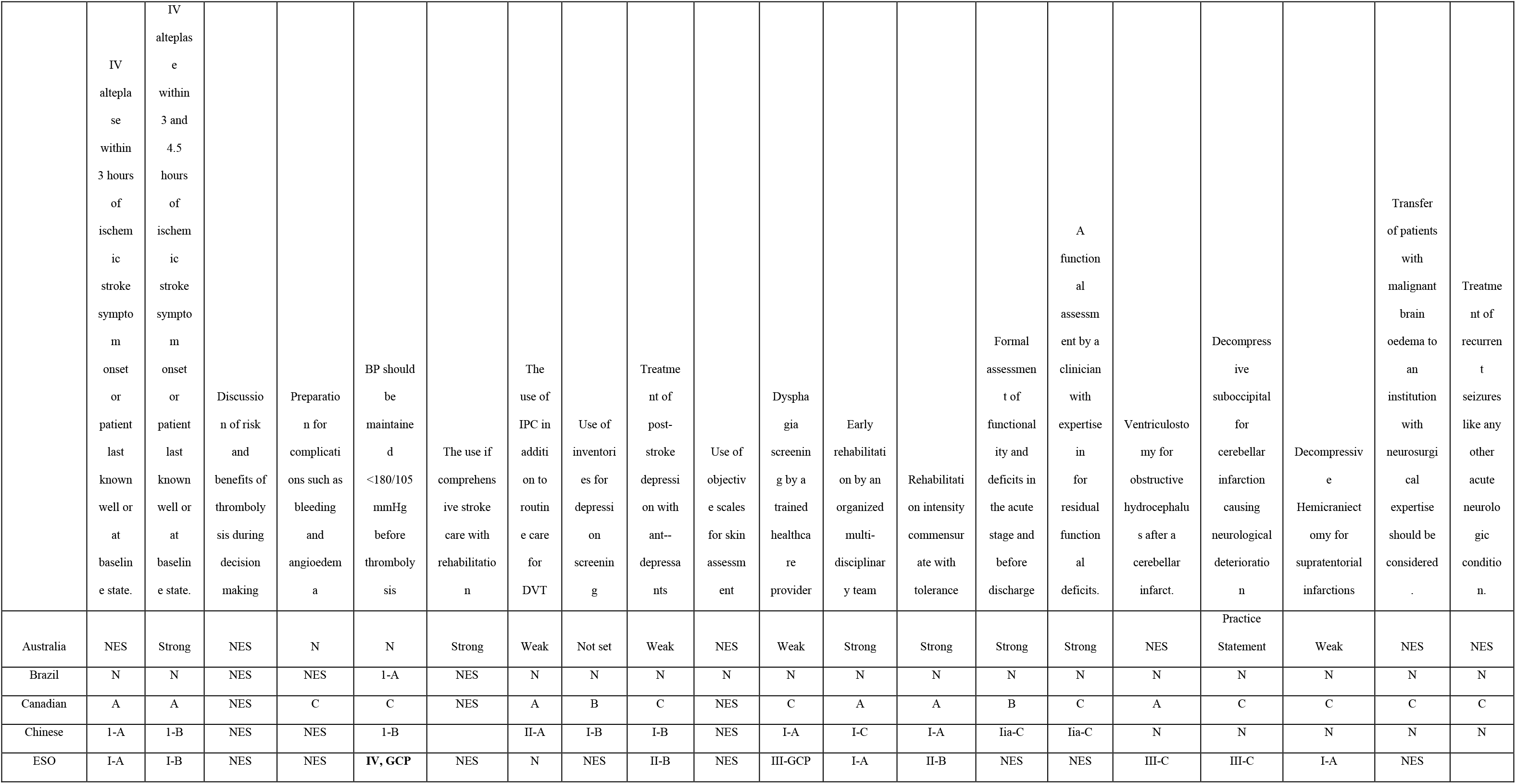
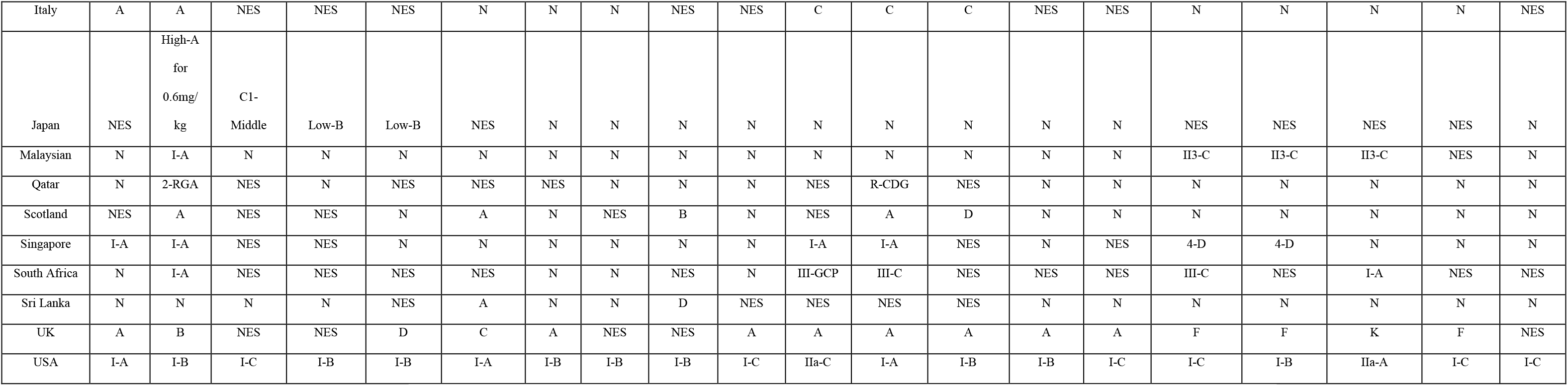
Summary of Recommendations in Various Stroke Guidelines

### Regionalization and Adaptation from other clinical guidelines

Most countries worldwide have no available published national guidelines. However, this does not translate to lack of systematic processes and workflows in the management of acute ischemic stroke. The European Stroke Organization has successfully implemented the ESO Stroke Guidelines which is being operationalized by countries in the European region (16). A unified approach is also being implemented in Australia and New Zealand as they both adapt the Australian Clinical Guideline for Stroke Management published in 2017 (27). More recently, the Middle East and North Africa Stroke and Interventional Neurotherapies Organization has also created a consolidated plan to manage stroke in the midst of the pandemic (28).

Most of the conceptualized guidelines have been adapted from existing ones, usually from high income countries (29). A systematic review comparing stroke clinical practice guidelines (CPGs) from low and high-income countries revealed that a degree of compromise in terms of the quality on the former (29). It is in this reason that in 2014, the World Stroke Organization conceived the *WSO Global Stroke Services Guideline and Action Plan* (30). This initiative aims to aid country-level health authorities to set-up or improve existing stroke frameworks to achieve high-quality, evidence-based recommendations and ensure that outcomes are measured to foster a milieu for continuous improvement (29).

### The need for grading recommendations

While countries have their own specific treatment recommendations, grading of evidence with the use of standardized systems are lacking. It is essential for guidelines to incorporate these as it ensures transparency and some level of confidence as these recommendations are translated into clinical practice (31). Various country-specific guidelines make use of their own grading systems in assessing the weight and level of evidence of the recommended guidelines (29). It is also essential for grading systems to be customized accordingly for low-income countries. Epidemiologists suggest the adaptation of internationally recognized approaches with efforts to integrate local evidence and weigh in appropriate resources (32).

### Clinical trials that changed the guidelines

In 1994, the AHA published the first clinical guideline on the management of acute ischemic stroke (33). While the efficacy of thrombolytic therapy was already being recognized then, it remained to be in the sidelines for safety concerns (34,35,37). With the encouraging results of the NINDs trial and the subsequent approval of alteplase by the US-FDA for systemic reperfusion, the AHA guidelines was updated, and it was also within this period that the Canadian Stroke Guidelines was conceived (36). Other major clinical guidelines from different parts of the world were also published subsequently.

With the aim of further improving stroke care, further modifications of then existing guidelines have been made. With the promising results of the ECASS3 trial, the time period for thrombolysis has been extended from three to 4.5 hours (38–39). The results of the J-ACT trial in 2006 has also resulted in the approval for use of the 0.6mg/kg dose of alteplase as mandated by the revised 2009 Japanese guidelines for stroke (18, 40)

The results of five clinical trials on endovascular therapy from 2012 to 2014 have also revolutionized the landscape of stroke management in the year 2015. The MR CLEAN, ESCAPE, EXTEND-IA, SWIFT-PRIME and the REVASCAT trials showed statistically significant improvement in clinical and radiologic outcomes after endovascular therapy (EVT) for large vessel occlusion (41–45). Clinical guidelines were revised so that patients within six hours from onset of symptoms were deemed eligible for EVT (12,14,26). A few years later, this time period was extended to 18-26 hours based on perfusion imaging parameters, as demonstrated by the DAWN and the DEFFUSE 3 trials (46, 47). Various clinical trials are still in the pipeline and are expected to make significant changes in existing guidelines worldwide in the future.

### Initial assessment

Guidelines included in the study are represented from all parts of the world including Asia, Australia, Europe, Africa and America (Table 2). Sections of clinical guidelines are subdivided into initial assessment, supportive treatment, reperfusion therapy, management of complications and rehabilitation. In most guidelines, pre-hospital and preventive strategies are usually included, but these are not discussed in this study. It is noted that some degree of variability in grading exists with the appraisal of different clinical guidelines.

In terms of initial assessment, there is unanimity in the clinical strategies that all patients suspected to have stroke should have neuroimaging urgently. This received the strongest recommendation among most of the countries with only ones from USA and Qatar, putting significant weight on aiming less than 20 minutes for it to be accomplished. High-income countries who have established facilities for endovascular thrombectomy also put the priority on neurovascular imaging. On the other hand, only a number of countries emphasize the use of scales for stroke severity assessment.

The importance of neuroimaging cannot be overemphasized in the management of acute stroke. While seamless processes to ensure efficiency in initial brain scanning have already been established in high-income countries, limitations in resources and logistics are still problematic most especially in rural areas of low to middle-income countries (48, 49). For example, a tertiary centre in India identified that the lack of neuroimaging facilities posed as one of the most important barriers for thrombolysis, with even the out of pocket cost for CT scan contributing to this limitation (50). It is also practical for other countries such as Sri Lanka, Serbia(51), South Africa and Malaysia not to put too much weight on neurovascular imaging as inaccessibility to neuro-interventionists and comprehensive stroke centres, as well as the high cost of treatment for this sophisticated procedure(51, 52), is still one of the identified problems in most developing countries (53). On the other hand, among countries in which reputable standard operating procedures for neuroimaging are already existent, aiming to shorter door to imaging times are being optimized, as trends to improved clinical outcomes have been observed (54).

There is also homogeneity among different countries in terms of what ancillary tests are to be performed during the hyperacute management of stroke. Serum blood glucose is being specified as an absolute test to be done prior to thrombolysis in some countries while in some, this is not explicitly identified. There is also unanimity among different countries that troponin, immune cell counts, and ECG should not be deterrents to timely thrombolysis. While obtaining baseline temperature is deemed significant in almost all clinical guidelines, less degree of weight is put in this parameter as opposed to blood glucose.

It has long been recognized that hypo and hyperglycaemia are known stroke masquerades (55). A study in 2015 among 80 consecutively recruited hypoglycemic patients revealed that 11% had stroke-like presentation with symptoms reversing within one hour of administration of intravenous dextrose (56). Furthermore, it is also essential that this parameter be recognized and corrected at an early stage as glycaemic aberrations in the peri-thrombolysis period may significantly impact on clinical outcomes (57). While deemed equally important, cardiac investigations should not preclude nor delay thrombolysis. It has been demonstrated in various studies that the presence of strain pattern, t-wave alterations, QT dispersion may be predictors of poor outcomes among stroke patients (58–60). Troponin is also essential to exclude the co-occurrence of AIS and acute myocardial infarction. A national registry including more than 800,000 patients with AIS identified that simultaneous occurrence of both only happens in 1.6% of the patients (61). While the incidence is significantly low, substantial increase in hospital mortality has been observed (61).

### General supportive care

There is heterogeneity in terms of supportive care among acute stroke patients with most clinical guidelines stressing moderate to strong recommendations on airway protection, correction of fluid imbalances and treatment of sources of hyperthermia and hypoglycaemia. Consensus for blood pressure targets are not uniform, with Caucasian guidelines emphasizing a threshold of 180/105 prior to thrombolysis while some Asian guidelines follow a higher target (19, 22).

It is essential that acute stroke units be organized in a manner that caters to the efficient provision of above-mentioned parameters as this has been positively associated with good outcomes such as reduction of mortality, length and cost of hospitalization as well as institutionalization (62, 63). This is particularly problematic in low to middle income countries because of concerns for costs, facilities and hospital staffing(64) Contrary to this, a recent prospective observational study in a tertiary hospital in South Africa demonstrated that despite the resource limitations, adaptation of the acute stroke response network which integrates organization of an acute stroke unit yields favourable thrombolysis outcomes at par to those observed in developed countries (65, 66).

Evidence proves that blood pressure optimization during thrombolysis results in good functional outcomes (67). Prospective and retrospective studies as well as clinical trials reveal that blood pressure during thrombolysis ranging from 140-160 reduced the odds of poor outcomes (68–69). To date, no studies have identified the most optimal blood pressure to achieve best outcomes post reperfusion therapy, however, clinical trial targets are set at 180/105, hence the parameters set in clinical guidelines (70).

### Thrombolysis and the management of medical and surgical complications

There is also agreement between different guidelines that thrombolytic therapy at a dose of 0.9mg/kg be instituted among eligible patients who arrive between three to 4.5 hours from the onset of symptoms. It is only the Japanese guideline which has approved of the use of the lower dose (0.6mg/kg). Also, only a few guidelines explicitly emphasize recommendations on the management of bleeding and angioedema after treatment. Neurosurgical recommendations for the management of malignant infarcts and obstructive hydrocephalous are also clearly defined in medium and high-income countries

Majority of the clinical trials which looked at the safety and efficacy of the low-dose alteplase was employed among Asians, specifically Japanese(71, 72). The favourable results of the J-ACT, ENCHANTED, THAWS trial support the Japanese recommendations (70,71,72,73). Aside from practical reasons of the lower cost from the reduced dose of alteplase (which usually just consumes 1 vial per dose), physiologic advantages such as lower levels of fibrinogen and Plasminogen Activator Inhibitor-1 (PAI-1) along with less marked genetic polymorphisms that induces a higher state of coagulation compared to Caucasians have also cited by Ueshima and colleagues (74). On the other hand, thrombolysis of patients with unclear onset of symptoms but with eligibility according the neuroimaging parameters of the WAKE-UP trial have also made the Australian and the AHA stroke guidelines recommend in favour of the later (75).

It is also interesting to note that of the guidelines reviewed, only three had explicitly stated recommendations on the management of thrombolysis-related complications such as bleeding and angioedema. More so, of the Asian countries included, only Japan had clear statements with this regard(76). It is equally important to address these limitations especially in resource-limited regions such as Asia and South America, where there is also a scarcity of stroke intensive care units (77, 78).

Encouraging results of various clinical trials for the management of malignant supra and infratentorial infarctions have been instrumental for the increase in confidence for guidelines to recommend these procedures especially for highly eligible patients. While this is of no question for countries with sufficient infrastructure and manpower, it has always been challenging for low- and middle-income countries. In sub-Saharan Africa, it has been previously identified that the ratio of neurosurgeon to population is as low 1: 64,000,000 (79). Furthermore, a study in 2015 on the economic losses attributed to neurosurgical diseases revealed that stroke was a major contributor to the three trillion macroeconomic deficits particularly in low income countries (80). It is therefore critical that guidelines be crafted according to individually available resources to ensure optimal implementability.

### Post-stroke rehabilitation

Stroke rehabilitation is another key component of stroke clinical guidelines. Majority put significant weight on early rehabilitation while moderate to weak strengths have been tagged for professional dysphagia assessment. The American, Australian and UK guidelines likewise put high premium on functional assessment while heterogeneity exists on integrating rehabilitation on comprehensive stroke care centre as well as the use of intermittent pneumatic compression for deep vein thrombosis. Majority of the guidelines have weak or no recommendations for depression screening and treatment, as well as regular skin assessment.

One of the aspects of stroke care that most clinicians fail to put attention into is post-acute rehabilitation. It is important for healthcare systems to adhere to posts-stroke rehabilitation guidelines as various studies have shown that compliance is positively correlated with good clinical outcomes (81–82). It has also been shown that low-cost rehabilitation (83) with focus on exercise-based and brain training interventions, in resource-deprived settings still translated to good clinical outcomes (84). Commensurate rehabilitation initiated within the first seven days of stroke has been shown to initiate complex neurobiological processes which is instrumental in early neurologic recovery as evidenced in various clinical trials (85, 86). Various clinical settings have also confirmed that post-stroke dysphagia results in aspiration pneumonia which further complicates hospital outcomes (88,89,90). Additionally, evidence-based practices for the prevention of deep vein thrombosis such as the use of IPC, should likewise be integrated as it may likewise impact on survival (91). Likewise, there should be increased vigilance for post-stroke depression among clinicians as it may occur in more than one third of stroke cases (91). The need to integrate this in clinical guidelines could not be over-emphasized especially in low-income to middle income countries due to its increasing prevalence (92, 93). Moreover, its impact on the disability-adjusted life-years lost is significantly greater in than in high income countries (94, 95)

### Ignored aspects of stroke care

The abundance of sophisticated techniques for stroke care has led clinicians to forget about the basic yet practical aspects of stroke management. It is noted that none of the stroke guidelines incorporate the use of basic immune biomarkers such as the neutrophil to lymphocyte ratio. In the advent of precision medicine nowadays, clinical practice is shifting towards accurate and specific disease characterization, as well as quantifying disease progression and response to therapy, for which biomarkers play critically important role (96). The neutrophil to lymphocyte ratio is a cheap, readily available and easy to interpret immune marker which may provide a diagnostic clue particularly for clinical outcomes post stroke (97–99).

While not mentioned in any of the clinical guidelines, the importance of ocular examination in stroke care should not be discounted. Fundus photography is an emerging tool which may assist in differentiating of stroke and TIA from other causes of neurologic deficits, particularly in the emergency setting (100). Retinal imaging, otherwise known as the ‘window to the brain’ may supplement neuroimaging particularly in providing insights for cerebrovascular neurodegenerative conditions (101). Lastly, it may also provide additional information for identifying stroke aetiology, especially that of complicated ones (102).

## Discussion

Stroke and post stroke complications culminating in massive health and economic impacts globally(103). Stroke occurs in a compromised vascular system. The risk factors associated with stroke ( both non-modifiable risk factors such as genetic, age, gender and modifiable risk factors such as hypertension, diabetes, high cholesterol, sedentary lifestyle, reduced fruits and vegetable intake, obesity, atrial fibrillation, poor air quality, smoking are linked with the build-up of low-grade chronic inflammation that perturbs the homeostasis of the vascular bed prior to the index vascular event such as acute stroke.

The index vascular event leads to a cascade of events that involve bioenergetic failure, disrupted cellular homeostasis, excitotoxicity, acidosis, damaged blood brain barrier and cell death very much akin to COVID-19 and brain involvement(104, 106). Contrary to the traditional belief that brain and immune systems are physically separate systems the neural and immune systems are intimately linked through SNS, HPA axis and also through glymphatic systems where bidirectional communication does occur regularly(107, 108).

There were 80.2 million (74.1 to 86.3) prevalent strokes globally in 2016 (109). Post stroke cognitive impairment has been reported over 50% ( which is still a gross underestimation) of stroke survivors with worsened disability and quality of life (110). Frequency of anxiety after stroke very high at 24.2% (21.5%-26.9%) by rating scales (111) with likely increased risk of further stroke and downward spiral from the PNI point of view. Post stroke depression (PSD) is reported at 18%-33% (112) ( gross under estimation again, see the comprehensive review on pathobiology of PSD Pascoe & Crewther (113)). Post Stroke Fatigue (PSF) is reported as one of the worst symptoms by 40% of the stroke survivors with prevalence of PSF vary from 25%-85% (114,115,117). Post Stroke Apathy(PSA) with a prevalence 34.6% and Central Post Stroke Pain ( CPSP) with a prevalence vary from 8%-55% (116, 117),can be added to long list of post stroke neurological complications with a similar psycho-neuroimmunological pathobiology to the PCNS as we elaborated in a series of recent publications (104–106)

We suggest the desperate need of systems biology approach to all these complications and conder the complete picture with a view to optimise the best immune response after the index event of acute stroke and re-visit the current guidelines as a matter of high priority. Such an approach will help the world to address one of the most disabling brain disorders affecting well over 80 million people with excellent value for money with current management approach and also the potential for individualized therapeutic and management avanues.

## Conclusion

Stroke management is a dynamic process which has evolved at a very fast pace over the past two decades. With the abundance of clinical trials in this field, it is possible that trends of management now may not be applicable in the future. Despite this, it is disappointing to see the lack of incorporation of easily accessible, low cost immune inflammatory prognostic markers such as NLR, or functional vision assessment at the bed side in any of the published stroke guidelines anywhere in the world. This is despite the fact that large number of publications and metanalyses supporting the role of immune inflammatory markers such as NLR in acute stroke as well as in the context of post stroke trajectory. It is therefore imperative for country-specific standard operating procedures to be updated constantly to fit to emerging needs with a systems biology-based approach. Implementability of clinical guidelines is anchored on evidence-based and well-appraised clinical guidelines which are customized according to available resources and to the beliefs of its end-users.

## Data Availability

data available on a reasonable request

